# Computer adaptive testing to assess impairing behavioral health problems in emergency department patients

**DOI:** 10.1101/2022.01.08.22268937

**Authors:** Lauren M. O’Reilly, Azhar I. Dalal, Serena Maag, Matthew T. Perry, Alex Card, Max B. Bohrer, Jackson Hamersly, Setarah Mohammed, Kelli Peterson, David G. Beiser, Robert D. Gibbons, Brian M. D’Onofrio, Paul I. Musey

## Abstract

**Study Objective:** To assess the prevalence of several impairing behavioral health domains in stable patients presenting to the emergency department (ED) through the use of computer adaptive testing and the association among the domains, standard assessments, and ED utilization.

**Methods:** This was a single-center observational study of 828 randomly selected adult patients presenting to the ED from May 2019 to March 2020. The main outcomes of interest were the self-administered, validated computer adaptive assessments of suicidality, depression, anxiety, PTSD, and substance abuse using computer adaptive testing, the CAT-MH™. We estimated the association among the CAT-MH™ subscales, standard assessments, and the number of ED visits in the year prior and 30 days after enrollment.

**Results:** The proportion of those who scored above the threshold of low/mild risk were: 24.1% (suicidality), 8.3% (depression), 16.5% (anxiety), 12.3% (PTSD), and 20.4% (substance use). While the CAT-MH™ subscales were highly correlated with other self-report assessments, they were not highly associated with standard ED assessments of suicidality. When examining associations with ED use, for example, individuals who had two or more ED visits in the prior year had 51% increased odds of being in the intermediate-high suicide risk category (95% Confidence Interval [CI], 1.03-2.23) compared to those with zero prior ED visits.

**Conclusion:** The CAT-MH™ can document the high prevalence of mental health conditions in the ED, which were missed by standard ED practices. Mental health problems are associated with ED utilization in a population of patients presenting largely for somatic complaints.

## Introduction

### Background

There are approximately 140 million visits to US Emergency Departments (ED) annually.^1^ Visits for psychiatric or substance abuse account for approximately 10% of these visits.^2^ Importantly, this represents a dramatic rise (approximately 30%) in the number of visits primarily for mental health complaints over the past two decades, which has outpaced the rise in overall ED visitation.^2-9^ ED service delivery costs associated with the most prevalent mental health and substance abuse diagnoses are in excess of $3.5 billion, with anxiety and depression accounting for $1.7 billion annually.^10^ The severity of mental health symptomology is positively correlated with rate of ED use.^11, 12^ Yet, most mental health problems are missed during ED encounters.^13-15^ In fact, 45% of patients who are seen in the ED for non-psychiatric primary complaints have comorbid psychiatric disorders, including major depression, general anxiety, substance abuse, and suicidal ideation.

### Importance

The barriers to standard true universal mental health screening in the ED are many and include the competing priorities of ED providers providing care to critically ill patients in busy and crowded ED environments, limited access to adequate mental health resources for consultation and outpatient care, as well as the training and attitudes of ED clinicians.^16, 17^ These same barriers have limited the approach to widespread mental health screening in the ED and represent a major missed opportunity to provide appropriate care as a significant proportion of ED patients presenting with non-psychiatric complaints have comorbid, unaddressed mental health issues as noted above. It is noteworthy that many patients who die by suicide often visit the ED or access the healthcare system shortly before their attempt for a non-psychiatric reason.18, 19

### Goals of this Investigation

In this study, we build and expand upon the work of Beiser et al. (2019) who demonstrated the feasibility of using a brief computer adaptive testing tool (the CAD-MDD^12^ and the CAT-DI^20^) for depression assessment in a broad sample of ED patients with non-psychiatric primary complaints. Our objectives in this study were three-fold. First, we aimed to document the prevalence of suicidality, depression, anxiety, post-traumatic stress disorder (PTSD), and substance abuse in an ED population. Second, we compared the results of the computer adaptive test-mental health (CAT-MH™) standardized screening to standard of care ED assessments for suicide, as well as to other validated self-report screening tools for anxiety and depression. Finally, we aimed to estimate the health care utilization of ED patients with mental health care problems.

## Methods

### Sample

This was a single-center prospective observational sample of 828 randomly selected non-consecutive patients presenting at a Midwestern emergency department (ED) over the course of 10 consecutive months. This study was approved by the Indiana University School of Medicine Institutional Review Board (protocol no.1904527368) and all participants gave their informed consent. Data collection began on May 24, 2019, and ceased on March 10, 2020 with the rapid increase of COVID-19 cases and resulting stay-at-home orders. Exclusion criteria for the study included: 1) age younger than 18 years, 2) being under involuntary detention, 3) lack of or impaired decisional capacity (e.g., active psychosis, dementia, developmental delay, intoxication), 4) hemodynamic instability according to the treating provider, 5) non-English speaking, and 6) those that may encounter issues with utilization review (e.g., prisoner, living out of the state). Therefore, the cross-section of included patients were relatively stable ED subjects without overt barriers to providing consent.

### Protocol

Trained research personnel used the ED tracking system within the electronic medical record (EMR) system (*Cerner Firstnet*) to screen for potentially eligible subjects presenting to the ED primarily during weekday daytime hours. To reduce sampling bias, research personnel began each screening shift by selecting a random digit between 0 and 9 using an online random number generator. All subjects on the ED census at that time with the last digit of their age matching the selected number were deemed eligible. This random selection process was repeated each time the list of eligible subjects was exhausted. The treating providers for screened subjects were approached to confirm eligibility criteria. Those subjects meeting all enrollment criteria were approached for informed consent. Participants then completed a multidimensional psychological screening using the self-administered CAT-MH™, which is described below. Additional data including medical comorbidity, past medical history, and chief complaint were collected and supplemented by EMR review. Socio-demographic data was collected through patient self-report. Self-reported past medical history was assessed by asking the participant to indicate if a doctor or another health care worker had diagnosed or treated the participant for 13 medical items in the past three years (see **Table 1**). Subsequently, each participant underwent a self-administered unidimensional screening for anxiety, depression, physical symptoms, and pain via a secure tablet.

**Table 1.** Frequency distribution of intake variables for entire sample and sample screening intermediate-high suicide risk on CAT-MH™

|  | Individuals consenting<br>for CAT-MH<sup>TM</sup><br>(N=794) | Individuals screened<br>intermediate-high CAT-MH<sup>TM</sup> suicide risk<br>(N=191) |
| --- | --- | --- |
| **Demographic Information** | N (%)<sup>a</sup> | N (%)<sup>b</sup> |
| Gender |  |  |
| Female | 492 (62.0) | 125 (65.5) |
| Male | 301 (37.9) | 66 (34.6) |
| Other | 1 (0.1) | 0 (0) |
| Ethnicity |  |  |
| Hispanic or Latino | 42 (5.3) | 11 (5.8) |
| Not Hispanic or Latino | 725 (91.3) | 174 (91.1) |
| Unknown or not reported | 27 (3.4) | 6 (3.1) |
| Race |  |  |
| White | 413 (52.0) | 105 (55.0) |
| Black or African American | 316 (39.8) | 65 (34.0) |
| Asian | 3 (0.4) | 0 (0) |
| American Indian/Alaska Native | 6 (0.8) | 2 (1.1) |
| Native Hawaiian or Other Pacific Islander | 0 (0) | 0 (0) |
| More than one race | 36 (4.5) | 15 (7.9) |
| Other | 20 (2.5) | 4 (2.1) |
| Marital Status |  |  |
| Single | 422 (53.2) | 101 (52.9) |
| Married | 224 (28.2) | 38 (19.9) |
| Separated | 21 (2.6) | 9 (4.7) |
| Divorced | 91 (11.5) | 31 (16.2) |
| Widowed | 36 (4.5) | 12 (6.3) |
| Education |  |  |
| Some high school | 126 (15.9) | 43 (22.5) |
| High school | 230 (29.0) | 39 (20.4) |
| GED | 42 (5.3) | 12 (6.3) |
| Some college | 210 (26.5) | 51 (26.7) |
| Graduate college | 149 (18.8) | 38 (19.9) |
| Graduate/professional school | 37 (4.7) | 8 (4.2) |
| Employment |  |  |
| Employed full-time | 318 (40.1) | 63 (33.0) |
| Employed part-time | 86 (10.8) | 26 (13.6) |
| Student and working | 19 (2.4) | 3 (1.6) |
| Student and not working | 19 (2.4) | 4 (2.1) |
| Homemaker | 19 (2.4) | 7 (3.7) |
| Unemployed | 99 (12.5) | 28 (14.7) |
| Retired | 80 (10.1) | 10 (5.2) |
| Disabled | 154 (19.4) | 50 (26.2) |
| Chief Complaint |  |  |
| Abdominal pain | 167 (21.0) | 45 (23.6) |
| Abnormal lab/test results | 3 (0.4) | 2 (1.1) |
| Abnormal vital signs (heart rate, blood pressure, etc.) | 14 (1.8) | 1 (0.5) |
| Allergic reaction | 3 (0.4) | 1 (0.5) |
| Anxiety | 3 (0.4) | 2 (1.1) |
| Chest pain | 91 (11.5) | 17 (8.9) |
| Ear, Nose, and Throat complaint | 28 (3.5) | 6 (3.1) |
| Eye complaint | 7 (0.9) | 0 (0) |
| Facial pain | 8 (1.0) | 2 (1.1) |
| Fever/chills/sepsis | 12 (1.5) | 0 (0) |
| Other musculoskeletal pain (back, extremity, etc.) | 105 (13.2) | 25 (13.1) |
| Gastrointestinal problem (vomiting, diarrhea, etc.) | 51 (6.4) | 14 (7.3) |
| Genitourinary problem (kidney complaints, etc.) | 14 (1.8) | 4 (2.1) |
| Headache | 26 (3.3) | 5 (2.6) |
| Medical screen/evaluation | 10 (1.3) | 3 (1.6) |
| Neurological complaint (seizure, focal weakness, etc.) | 18 (2.3) | 4 (2.1) |
| Obstetric-Gynecological complaint | 21 (2.6) | 7 (3.7) |
| Post-operative complication | 4 (0.5) | 2 (1.1) |
| Respiratory problem (cough, asthma, shortness of breath) | 78 (9.8) | 15 (7.9) |
| Sickle cell crisis | 11 (1.4) | 1 (0.5) |
| Skin/soft tissue infection or complaint | 22 (2.8) | 6 (3.1) |
| Substance use | 1 (0.1) | 1 (0.5) |
| Trauma (motor vehicle accident, fall, assault, etc.) | 34 (4.3) | 5 (2.6) |
| Weakness/fatigue and dizziness | 44 (5.5) | 14 (7.3) |
| Miscellaneous | 16 (2.0) | 8 (4.2) |
| Invalid | 2 (0.3) | 1 (0.5) |
| Missing | 1 (0.1) | 0 (0) |
| **Health History Information** |  |  |
| Currently have a primary care physician | 580 (73.0) | 136 (71.2) |
| History of: |  |  |
| Asthma, emphysema, chronic bronchitis | 251 (31.6) | 75 (39.3) |
| High blood pressure or hypertension | 341 (42.9) | 82 (42.9) |
| High blood sugar or diabetes | 175 (22.0) | 46 (24.1) |
| Arthritis or rheumatism | 226 (28.3) | 63 (33.3) |
| Heart attack, angina, heart failure, or other types of heart disease | 140 (17.6) | 36 (18.9) |
| Stroke, seizures, Parkinson's disease, or another neurological condition | 102 (12.8) | 40 (20.9) |
| Liver disease | 40 (5.0) | 12 (6.3) |
| Kidney or renal disease | 82 (10.3) | 27 (14.4) |
| Cancer diagnosed or treated in the last 3 years | 58 (7.3) | 12 (6.3) |
| Anxiety | 323 (40.7) | 136 (71.2) |
| Depression | 307 (38.7) | 141 (73.8) |
| PTSD | 129 (16.2) | 67 (35.1) |
| Suicidal thoughts or attempt | 58 (7.3) | 45 (23.6) |
| Current use of: |  |  |
| Tobacco | 269 (33.9) | 88 (46.1) |
| Alcohol $\geq 4$ times/week | 99 (12.5) | 28 (14.7) |
| Marijuana | 163 (20.5) | 73 (38.2) |
| Cocaine | 13 (1.6) | 5 (2.6) |
| Heroin | 3 (0.4) | 1 (0.5) |
| Other | 13 (1.6) | 7 (3.7) |
| **Symptom Questionnaire Information** | **N (%)** | **N (%)<sup>b</sup>** |
| GAD-7 (Generalized Anxiety Disorder) |  |  |
| Minimal (0-4) | 454 (57.2) | 22 (11.5) |
| Mild (5-9) | 184 (23.2) | 60 (31.4) |
| Moderate (10-14) | 73 (9.2) | 40 (20.9) |
| Severe ($\geq 15$) | 83 (10.6) | 69 (36.1) |
| PHQ-8 (Depression) |  |  |
| Minimal (0-4) | 398 (50.1) | 15 (7.9) |
| Mild (5-9) | 188 (23.7) | 33 (17.3) |
| Moderate (10-14) | 118 (14.9) | 61 (31.9) |
| Moderately severe (15-19) | 58 (7.3) | 51 (26.7) |
| Severe ($\geq 20$) | 32 (4.0) | 31 (16.2) |
| PCS-13 (Pain Catastrophizing) |  |  |
| None/Mild (0-29) | 705 (88.8) | 124 (64.9) |
| Clinically significant ($\geq 30$) | 89 (11.2) | 67 (35.1) |
| PHQ-15 (Somatic Symptoms) |  |  |
| Very Low (0-4) | 135 (19.3)<sup>c</sup> | 10 (6.0)<sup>d</sup> |
| Low (5-9) | 236 (33.8)<sup>c</sup> | 35 (21.1)<sup>d</sup> |
| Medium (10-14) | 195 (27.9)<sup>c</sup> | 51 (30.7)<sup>d</sup> |
| High ($\geq 15$) | 133 (19.0)<sup>c</sup> | 70 (42.1)<sup>d</sup> |
| VAS Suspected suicide within next 30 days | M (SD) (range)<br>5.0 (8.9) (0-74) | M (SD) (range)<br>6.3 (11.3) (0-74) |
| **Admission Information** | M (SD) (range) | M (SD) (range) |
| Number of ED visits within 12 months prior to enrollment date | 2.1 (4.7) (0-76) | 3.3 (7.6) (0-76) |
| Number of repeated ED visits in 30 days since enrollment date | 0.3 (0.8) (0-9) | 0.5 (1.1) (0-7) |
| <sup>a</sup>Based on 794 unique individuals. Rounded to nearest tenth; may not equal 100%. <sup>b</sup>Based on 191 individuals. <sup>c</sup>Based on 699 unique individuals. <sup>d</sup>Based on 166 individuals. |  |  |

Health care utilization data was obtained through the EMR and the Indiana Network for Patient Care (INPC) managed by the Indiana Health Information Exchange, the nation’s largest inter-organizational clinical data repository,^21^ to collect data regarding disposition (admit or discharge), medical history, discharge diagnoses, and ED utilization in the 12 months before and after enrollment. Note that while we intended to examine ED utilization 12 months before and after enrollment, we limited our analyses to 12 months prior and 30 days after enrollment. Research has demonstrated that the COVID-19 pandemic greatly affected health care use, particularly in EDs,^22^ which would have systematically biased our long-term follow-up data.

During the study period, standard of care universal suicide screening took place at the enrollment site ED for individuals not presenting with a chief complaint of suicidal ideation. The standard of care consisted of three questions closely aligned with the Columbia-Suicide Severity Rating Scale (C-SSRS): 1) “Have you wished you were dead or wished you could go to sleep and not wake up?”; 2) “Have you had thoughts of harming yourself?”; 3) “In the last three months have you done anything, started to do anything or proposed to anything to end your life?” Individuals answering “yes” to number two were asked the remaining three questions of the suicidal ideation section of the C-SSRS and the attending physician was notified. Additionally, the patient’s provider was asked to quantify their concern for a future suicide attempt by marking a 100-millimeter visual analog scale (VAS) in response to the question: “What is your level of suspicion that this patient will attempt suicide in the next 30 days?”. Finally, the CAT-MH™, as described below, also screened for suicide risk. Treating clinicians were informed of anyone who screened in the high-risk category for suicide using this tool.

### Computer Adaptive Testing with the CAT-MH™

The CAT-MH™ ^20, 23^ is a suite of mental health computerized adaptive tests (CATs), that can be self-administered via any internet capable device (computer, tablet, smartphone), either in the clinic, ED, or remotely via a secure patient portal. The CAT-MH™ is unique in that unlike traditional CATs developed for educational assessments that are based on unidimensional item response theory (IRT) and are only appropriate for simple constructs such as mathematical ability, the CAT-MH™ is based on multidimensional item response theory and is suitable for the measurement of complex constructs such as depression, anxiety, and suicidality. For a given construct (e.g., depression) the CAT-MH™ draws symptom items from a large bank of potential items that completely covers the latent variable of interest from the lowest to the highest levels of severity. The CAT begins by selecting an item from the middle of the severity continuum. Based on the item response, typically on a 5-point Likert scale, a provisional severity score and its uncertainty are computed. Based on that score, the next maximally informative item is selected and administered. Based on the response to the second item, the severity score and uncertainty are recomputed. The process continues until the uncertainty in the estimated score falls below 5 points on a 100-point scale.

In our study we included CATs for suicidality (CAT-SS^24^), depression (CAT-DI^20^), anxiety (CAT-ANX^25^), PTSD (CAT-PTSD - an adaptive version of the 20-item PCL-5^26^), and substance use disorder (CAT-SUD^27^). The CAT-SUD also determines frequency of use of alcohol, sedatives/hypnotics, opioid analgesics, heroin/methadone, and cocaine/amphetamines. The suicide scale in the CAT-MH™ (the CAT-SS) was validated in two university hospital EDs,^24^ and the depression test (CAT-DI) has been validated for use in EDs.^12^

### Non-Adaptive Screening Tools

We administered four additional validated self-administered screening tools. (1) The Generalized Anxiety Disorder 7-item scale (GAD-7) is a rapid screening tool for the presence of clinically significant anxiety.^28-31^ The scores range from 0-21, with cut points of 5, 10, and 15 corresponding to mild, moderate, and severe anxiety, respectively. (2) The Patient Health Questionnaire 8 item scale (PHQ-8) is a rapid screening tool for the presence of clinically significant depression.^31-37^ The scores range from 0-24, with cut points of 5, 10, 15, and 20 corresponding to mild, moderate, moderately severe, and severe depression, respectively. (3) The Patient Health Questionnaire-15 (PHQ-15) is used to detect participants at risk for somatoform disorders;^38, 39^ it has cutoffs of 5, 10, and 15, corresponding to low, medium, and high somatic symptom severity. (4) The Pain Catastrophizing Score (PCS) is a 13-item self-administered tool used to quantify a person’s thoughts and feelings when they have experienced pain. ^40, 41^ The maximum total score is 52 and total scores ≥ 30 are considered clinically significant.^40^

### Analyses

We documented the percentage of patients endorsing different categories (e.g., normal, mild, moderate, severe) of the five domains assessed by the CAT-MH™. We present this for all the patients, as well as a subgroup of patients who scored in the intermediate to high categories of suicidal risk. We then compared the identification of patients at risk for suicide using the CAT-MH™ with the standard ED procedures. To further examine the assessment of mental health and substance problems via the CAT-MH™, we correlated the CAT-MH™ scores with clinician-reported suspected suicide and self-reported measures.

To determine the association between the number of ED visits in the year prior to enrollment and CAT-MH™ subscales, we conducted logistic regression analyses. The predictors, number of ED visits within the prior year, were dummy coded into three categories: 1) zero visits in the year prior, which served as the reference category, 2) one visit prior, and 3) two or more visits prior. We analyzed each CAT-MH™ subscale separately as a binary outcome, dichotomized suicide and substance use into low risk (0) and intermediate-high risk (1), depression and anxiety into normal/mild (0) and moderate/severe (1), and PTSD into no evidence (0) and possible/highly likely (1). We included the following demographic covariates: age, gender, ethnicity, race, marital status, and employment.

In order to examine the association between the CAT-MH™ subscales and subsequent ED visits within 30 days of ED enrollment date, we conducted logistic regression analyses adjusting for demographics. The predictors were each of the categorical risk levels for the CAT-MH™ subscales, which we dummy coded such that the lowest level of risk served as the reference category. Given the reduced sample size in the high-risk categories, we combined all the categories above low risk. For depression and anxiety subscales, mild was also included in the reference category. We dichotomized the outcome into not present (0) or any ED visit (1) within 30 days.

### Sensitivity Analyses

We conducted three sensitivity analyses to examine the potential impact of analytical decisions on the results and interpretation. First, we predicted the continuous CAT-MH™ subscales from ED visits in the year prior to enrollment. We included an unstandardized and standardized outcome to examine the magnitude of the effects in terms of a single point increase on the 100-point scale and in terms of standard deviation units, respectively. Second, when assessing the associations with mental health problems with subsequent ED visits, we utilized continuous, standardized CAT-MH™ subscales rather than utilizing categorical CAT-MH™ predictors. Third, we conducted ordinal logistic regression to predict ED visits within 30 days after enrollment to examine the association with each CAT-MH™ category.

## Results

In the study, 1,854 patients were screened and approached for enrollment. Of those subjects who were approached for enrollment in the study, 828 (44.7%) underwent informed consent and were enrolled. Of those who consented, 97.3% (n=806) completed the CAT-MH™. Reasons for lack of completion in the remaining 22 subjects included study withdrawal, interruptions related to clinical care such as procedures, and completion times over one hour. In order to conduct a complete case analysis, we dropped an additional 12 individuals with the following missing indicators: demographic variables, use of ED care within 30 days post-discharge, and the PTSD subscale on the CAT-MH™. We also dropped one individual with an extreme outlier of ED care (i.e., 75 visits) within 30 days post-discharge and two individuals with invalid ages. The final analytic sample consisted of 794 individuals. See **Supplementary Figure 1** for the data flowchart. The median time for CAT-MH™ completion for the five domains was 9.7 minutes (IQR, 5.6 minutes).

**Table 1** presents the overall breakdown of our population including frequency distribution of demographic, chief complaint, health history, suicide, admission, and questionnaire information of the subjects included in the sample. The average age was 43.7 years (Standard Deviation [SD], 16.3). Approximately 62% of the sample identified as female gender, 5% identified as Hispanic or Latino ethnicity, 51% identified as White/Caucasian, while 39% were Black/African American, 53% were single, 84% had a high school degree/GED or above, and 40% were employed full-time. The three most common chief complaints when participants presented in the ED were abdominal pain (21.0%), musculoskeletal pain (e.g., back, extremity, etc.) (13.2%), and chest pain (11.5%). The three most common health problems of which participants had a history included high blood pressure or hypertension (42.9%), anxiety (40.7%,), and depression (38.7%).

The mean score of the GAD-7 was 5.3 (SD, 5.9), PHQ-8 was 6.4 (SD, 5.8), PCS-13 was 10.7 (SD, 12.8), and PHQ-15 was 9.6 (SD, 5.6). The average number of ED visits to the Midwestern healthcare network in 12 months prior to the participant’s enrollment date was 2.1 (SD, 4.7) and within 30 days since the participant enrollment date was 0.3 (SD, 0.8).

Suicide risk severity was also assessed by the CAT-MH™ and the right-hand column in Table 1 presents the frequency distribution of demographic variables, chief complaint, medical history, self-report questionnaires, and admission information for those who screened intermediate-high-risk on the CAT-MH™ suicide severity rating (n=191). The frequencies of demographic, chief compliant, and medical information were similarly distributed for individuals who screened intermediate-high suicide risk as for the entire sample. Individuals who screened intermediate-high suicide risk had a higher proportion of those who endorsed history of PTSD or suicidal ideation, as well as a higher mean of prior ED visits within the past year, as compared to the entire sample.

**Table 2** presents the frequency distribution of the CAT-MH™ subscales defined both categorically and continuously. The proportion of those who scored above the threshold of low risk/no evidence group for the following subscales were: 1) 24.1% for suicide, 2) 12.3% for PTSD, and 3) 20.4% for substance use. The proportion of those who scored above the threshold of normal/mild risk group for the following subscales were: 1) 8.3% for depression, and 2) 16.5% for anxiety. Across all categories, approximately 31.9% (n=253) of the sample screened above the threshold for at least one CAT-MH™ subscale and 20.7% (n=164) for two or more CAT-MH™ subscales. See **Supplementary Table 1** for the frequency distribution of use and abuse of specific substances.

**Table 2.**
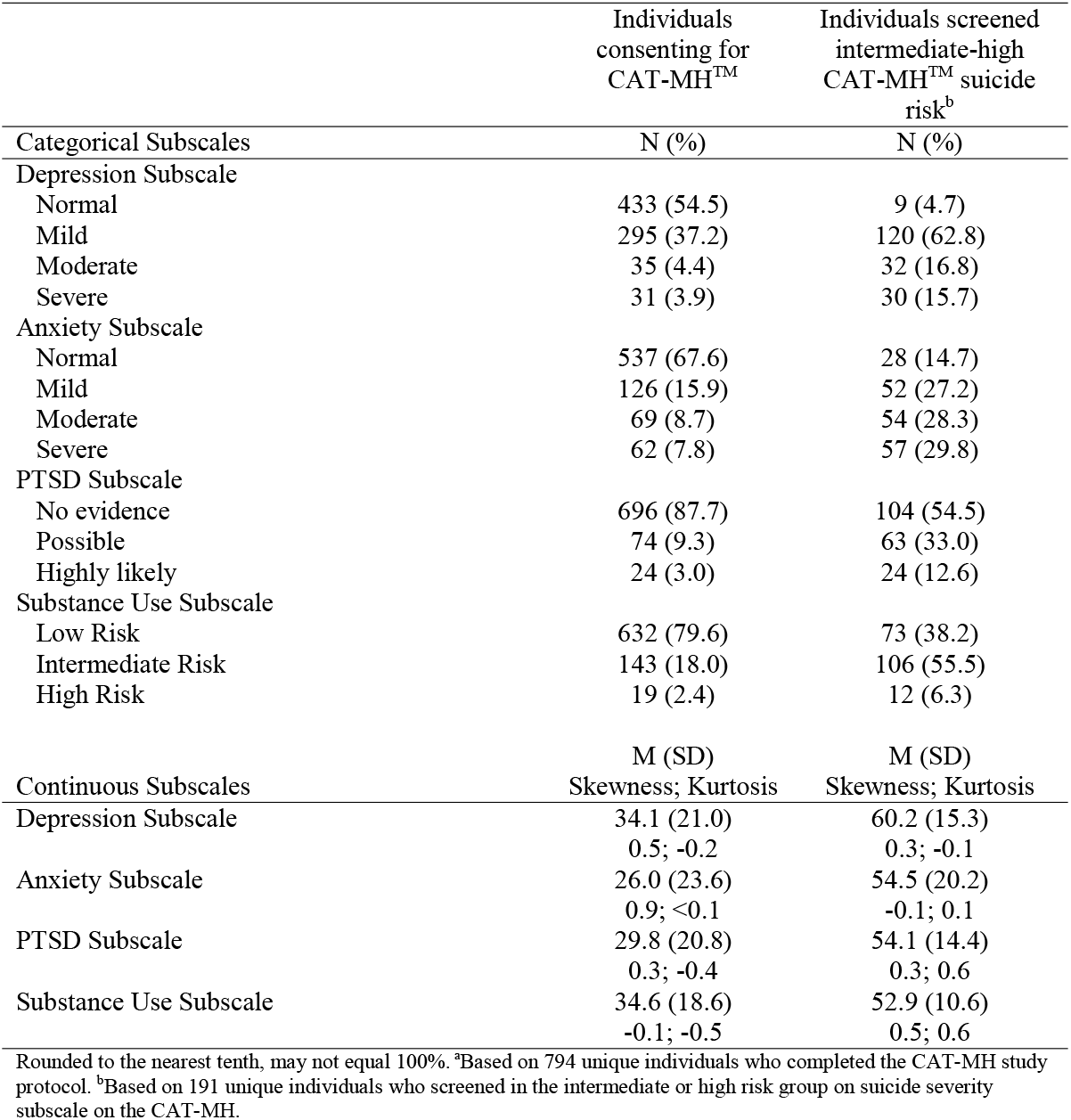
Frequency distribution of CAT-MH™ variables for entire sample and those who screened intermediate-high suicide risk on CAT-MH™

|  | Individuals<br>consenting for<br>CAT-MH<sup>TM</sup> | Individuals screened<br>intermediate-high<br>CAT-MH<sup>TM</sup> suicide<br>risk<sup>b</sup> |
| --- | --- | --- |
| Categorical Subscales | N (%) | N (%) |
| Depression Subscale |  |  |
| Normal | 433 (54.5) | 9 (4.7) |
| Mild | 295 (37.2) | 120 (62.8) |
| Moderate | 35 (4.4) | 32 (16.8) |
| Severe | 31 (3.9) | 30 (15.7) |
| Anxiety Subscale |  |  |
| Normal | 537 (67.6) | 28 (14.7) |
| Mild | 126 (15.9) | 52 (27.2) |
| Moderate | 69 (8.7) | 54 (28.3) |
| Severe | 62 (7.8) | 57 (29.8) |
| PTSD Subscale |  |  |
| No evidence | 696 (87.7) | 104 (54.5) |
| Possible | 74 (9.3) | 63 (33.0) |
| Highly likely | 24 (3.0) | 24 (12.6) |
| Substance Use Subscale |  |  |
| Low Risk | 632 (79.6) | 73 (38.2) |
| Intermediate Risk | 143 (18.0) | 106 (55.5) |
| High Risk | 19 (2.4) | 12 (6.3) |
|  | M (SD) | M (SD) |
| Continuous Subscales | Skewness; Kurtosis | Skewness; Kurtosis |
| Depression Subscale | 34.1 (21.0) | 60.2 (15.3) |
|  | 0.5; -0.2 | 0.3; -0.1 |
| Anxiety Subscale | 26.0 (23.6) | 54.5 (20.2) |
|  | 0.9; <0.1 | -0.1; 0.1 |
| PTSD Subscale | 29.8 (20.8) | 54.1 (14.4) |
|  | 0.3; -0.4 | 0.3; 0.6 |
| Substance Use Subscale | 34.6 (18.6) | 52.9 (10.6) |
|  | -0.1; -0.5 | 0.5; 0.6 |

Rounded to the nearest tenth, may not equal 100%. <sup>a</sup>Based on 794 unique individuals who completed the CAT-MH study protocol. <sup>b</sup>Based on 191 unique individuals who screened in the intermediate or high risk group on suicide severity subscale on the CAT-MH.

To examine the overlap between suicide screens, **Table 3** presents the cross-tabulation between the CAT-MH™ positive suicide screen (i.e., those in the high-risk suicide severity category) and the ED’s standard of care. Ten individuals screened positive on the CAT-MH™ suicide screen but only three of these screened positive by the standard of care ED screen. Thus, seven individuals were not identified via the standard of care screening. All seven identified participants were in the high-risk category on the CAT-MH™ suicide subscale. **Table 4** presents the Pearson correlations between the CAT-MH™ subscales and the clinician-rated suspected suicide rating and self-reported questionnaires. Each CAT-MH™ subscale, including suicidality, was weakly correlated with the clinician rating of suspected suicide, and was strongly correlated with the self-reported questionnaires indexing of anxiety, depression, pain catastrophizing, and physical symptoms.

**Table 3.** Cross-tabulation between suicide screen in ED and CAT-MH

|  | CAT-MH Suicide Screen (N, %) |  |
| --- | --- | --- |
|  | Negative | Positive<sup>1</sup> |
| ED Standard Care Suicide Screening |  |  |
| Positive | 0 (0) | 3 (0.38) |
| Negative | 760 (95.72) | 7 (1.01) |
| Not Recorded | 23 (2.90) | 0 (0) |

Derived from 794 unique individuals. CAT-MH suicide screen based on suicide severity. <sup>1</sup> Individuals who screened positive were in the high-risk suicide severity group.

**Table 4.**
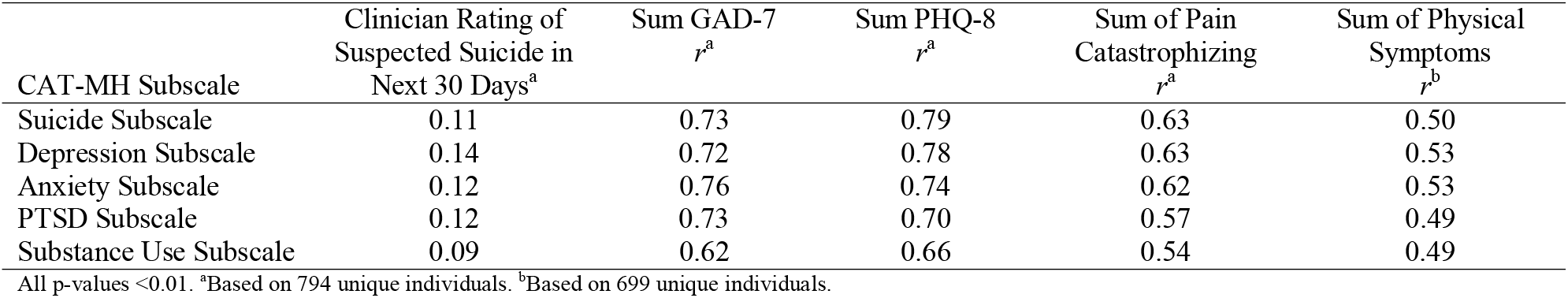
Pearson correlation between CAT-MH subscales, clinician-reported suicidality and other self-report measures

| CAT-MH Subscale | Clinician Rating of<br>Suspected Suicide in<br>Next 30 Days<sup>a</sup> | Sum GAD-7<br>*r*<sup>a</sup> | Sum PHQ-8<br>*r*<sup>a</sup> | Sum of Pain<br>Catastrophizing<br>*r*<sup>a</sup> | Sum of Physical<br>Symptoms<br>*r*<sup>b</sup> |
| --- | --- | --- | --- | --- | --- |
| Suicide Subscale | 0.11 | 0.73 | 0.79 | 0.63 | 0.50 |
| Depression Subscale | 0.14 | 0.72 | 0.78 | 0.63 | 0.53 |
| Anxiety Subscale | 0.12 | 0.76 | 0.74 | 0.62 | 0.53 |
| PTSD Subscale | 0.12 | 0.73 | 0.70 | 0.57 | 0.49 |
| Substance Use Subscale | 0.09 | 0.62 | 0.66 | 0.54 | 0.49 |

All p-values <0.01. <sup>a</sup>Based on 794 unique individuals. <sup>b</sup>Based on 699 unique individuals.

**Table 5** presents the prediction of dichotomous CAT-MH™ subscales from the number of ED visits in the year prior to enrollment. Individuals who had two or more ED visits in the prior year had a 51% increase in the odds of being in the intermediate-high suicide risk category (OR=1.51, 95% Confidence Interval [CI], 1.03-2.23) compared to those with zero prior ED visits. Odds ratios for the other CAT-MH™ subscales were the following: depression (OR, 1.61 [0.88-2.93]), anxiety (OR, 1.47 [0.94-2.29]), PTSD (OR, 2.49 [1.47-4.20]), and substance use disorder (OR, 1.55 [1.02-2.35]). Individuals with one visit to the ED in the prior year were not more likely to score higher on any of the CAT-MH™ subscales compared to those with zero visits. **Table 6** presents the prediction of dichotomous ED visits (not present/any) within 30 days from CAT-MH™ subscales. Individuals who scored in the intermediate-high-suicide risk group had 71% greater odds of an ED visit within 30 days after their enrollment compared to those who scored as low risk (OR=1.71, 95% CI, 1.16, 2.53). The associations with the other domains assessed by the CAT-MH™ were not statistically significant and the confidence intervals around the estimates were quite wide.

**Table 5.** Prediction of CAT-MH from prior year ED visits using logistic regression

| Visits ED year prior | Suicide Subscale<br>OR (95% CI; p-value) | Depression Subscale<br>OR (95% CI; p-value) | Anxiety Subscale<br>OR (95% CI; p-value) | PTSD Subscale<br>OR (95% CI; p-value) | Substance Use Subscale<br>OR (95% CI; p-value) |
| --- | --- | --- | --- | --- | --- |
| 0 | REF | REF | REF | REF | REF |
| 1 | 0.85 (0.53-1.37; p=0.50) | 1.03 (0.49-2.19; p=0.94) | 1.02 (0.60-1.76; p=0.93) | 1.54 (0.82-2.89; p=0.18) | 1.14 (0.69-1.86; p=0.62) |
| ≥2 | 1.51 (1.03-2.23; p=0.04) | 1.61 (0.88-2.93; p=0.12) | 1.47 (0.94-2.29; p=0.09) | 2.49 (1.47-4.20; p<0.001) | 1.55 (1.02-2.35; p=0.04) |

Derived from 794 unique individuals. Demographic variables included are age, gender, ethnicity, race, marital status, and employment. Each CAT-MH subscale was dichotomized: suicide subscale 0=low risk, 1=intermediate-high risk; depression subscale 0=normal-mild; 1=moderate-severe risk; anxiety subscale 0=normal-mild; 1=moderate-severe; PTSD subscale 0=no evidence, 1=possible-highly likely; substance use subscale 0=low risk, 1=intermediate-high risk.

**Table 6.** Prediction of ED visits within 30 days using logistic regression with combined categorical predictors

|  | OR (95% CI; p-value) |
| --- | --- |
| Suicide |  |
| Low Risk | REF |
| Intermediate-High Risk | 1.71 (1.16-2.53; p=0.01) |
| Depression |  |
| Normal-Mild | REF |
| Moderate-Severe | 1.06 (0.58-1.94; p=0.86) |
| Anxiety |  |
| Normal-Mild | REF |
| Moderate-Severe | 1.20 (0.77-1.89; p=0.43) |
| PTSD |  |
| No Evidence | REF |
| Possible-Highly Likely | 1.13 (0.67-1.89; p=0.65) |
| Substance Use |  |
| Low Risk | REF |
| Intermediate-High Risk | 0.95 (0.62-1.47; p=0.83) |

Derived from 794 unique individuals. ED visits dichotomized into not present/any (0/1). Demographic variables included are age, gender, ethnicity, race, marital status, and employment.

### Sensitivity Analyses

First, we predicted continuous CAT-MH™ subscales from visits to the ED within the 12 months prior to enrollment. The pattern of results was similar to the main analyses. For example, patients who visited the ED two or more times in the previous year scored 4.6 points higher on the continuous measure of suicide, which was a 0.25 standard deviation difference (**Supplementary Table 2**). Second, we predicted ED admissions 30 days after enrollment from standardized continuous CAT-MH™ subscales. The trend of results was similar to the main analyses (**Supplementary Table 3**). For suicide, each standard deviation increase in the suicide subscale was associated with a 19% increased odds of ED admission. Third, when conducting ordinal logistic regression to predict ED visits within 30 days after enrollment, the results followed a similar pattern as the main analyses, although the estimates were less statistically precise (**Supplementary Table 4**).

## Discussion

This prospective observational study represents the most complete use of the validated computer adaptive tool (CAT-MH™) in the ED environment for multidomain mental health screening in a broad random sample of patients presenting to a large urban emergency department. According to the results of the CAT-MH™ screening, approximately one-third of the sample population (31.9%) had at least one high CAT-MH™ subscale and 20% of the sample had at least two high subscales scores. For example, 16.5% and 8.3% screened positive for moderate/severe anxiety and depression, respectively. The prevalence of clinically significant depression in this population (8.3%) by CAT-MH™ screening is similar to the prevalence of moderate to severe depression noted (7%) in another ED sample using the CAT-MH™.^42^ Our population was primarily comprised of patients who presented without psychiatric complaint, and we know that a significant proportion of ED patients who are evaluated for non-psychiatric complaints (e.g., chest pain and abdominal pain) have comorbid mental health problems, which often remain undiagnosed during ED encounters.^13-15, 43^

Estimates suggest that approximately 1% of all US ED visits are related to suicidal ideation.^9^ In our enrolled sample, three participants screened positive for suicidal ideation according to standard ED screening procedures as described above. While the CAT-MH™ correctly identified each of these individuals and categorized them as high risk, this tool detected seven additional high-risk individuals not flagged by standard of care screening. Notably, clinician report of suspected suicide was only very weakly correlated (r=0.11) with the CAT-MH™ index of suicidality. It is possible that patients felt more at ease disclosing their feelings via the tablet administered CAT-MH™ rather than directly to the nurse at check in or during the evaluation by the doctor. The results illustrate that universal screening protocols are minimally time-intensive (i.e., the median response time was roughly 2 minutes per each of the five included domains) and effective at identifying patients at high risk who would have otherwise been missed due to the non-psychiatric nature of their chief complaint.^7, 44^ This is important, as a significantly higher proportion of those who screened intermediate to high suicide risk via the CAT-MH™ also self-reported more problems with anxiety, depression, PTSD, and substance use compared to the rest of the sample.

One of the aims of our study was to examine the relationship between mental illness and ED recidivism as mental illness has been shown to be a strong predictor of frequent ED use.^11, 12, 45^ In our study, compared to those with no visits in the 12 months prior to enrollment, those with two or more ED visits were significantly more likely to score in the high ranges for suicide risk, PTSD, and substance use on the CAT-MH™. With regard to subsequent ED use in the 30 days post-enrollment, suicide risk (intermediate-high on the CAT-MH™) was associated with an increased likelihood of repeat ED utilization. Taken together, these results are consistent with prior literature showing that approximately 30% to 50% of these patients with frequent ED utilization have a mental health or substance use disorder.^11, 46-49^

There are several important limitations to mention. We attempted to obtain a random sample of ED patients within our inclusion criteria with the aid of a random number generator in addition to adding evening and weekend screening, but this is only quasi-randomization. Additionally, since this study depended upon voluntary participation, it is unclear if those with underlying psychological conditions were more or less likely to volunteer to participate given either a willingness to disclose and seek help or concern regarding persistent mental health stigmas. In any case, these represent a form of selection bias which may actually underestimate the magnitude of impairing behavioral health conditions. With regards to the chief complaint, the overwhelming majority of complaints were somatic in nature. While we did not overtly seek to exclude those with mental health presentations (complaints of anxiety and depression), these patients are often triaged to the psychiatric observation rooms for close behavioral assessment, which would have effectively excluded them until they were moved out of those areas. As such, the results of this study apply primarily to patients receiving care in the ED for non-psychiatric complaints. Finally, our overall enrollment goals (1,000 patients) and ED recidivism targets (12 months) were curtailed by the institution of COVID-19 precautions and stay-at-home orders. We had planned to collect and analyze ED recidivism data for the 12 months post enrollment, but due to the extraordinary environmental and societal pressures applied by the broad stay-at-home orders during the COVID-19 pandemic^22^, we believed this would confound these data. Nonetheless, it is clear that there is a strong association between mental health issues and ED recidivism, which may benefit from screening tools such as the CAT-MH™.

In the study we were able to demonstrate the use of the validated multidimensional CAT-MH™ to document the high rate of impairing mental health conditions and their associated ED utilization in a population of ED patients presenting largely for somatic complaints. Furthermore, in this population the CAT-MH™ performed superior to standard of care suicide screening in identifying more than three times the number of individuals at high risk for suicide. Future work should focus on assessing the ability of the multidomain CAT-MH™ in universal screening.

## Data Availability

All data produced in the present study are available upon reasonable request to the authors

## Supplementary Material

**Supplementary Figure 1.**
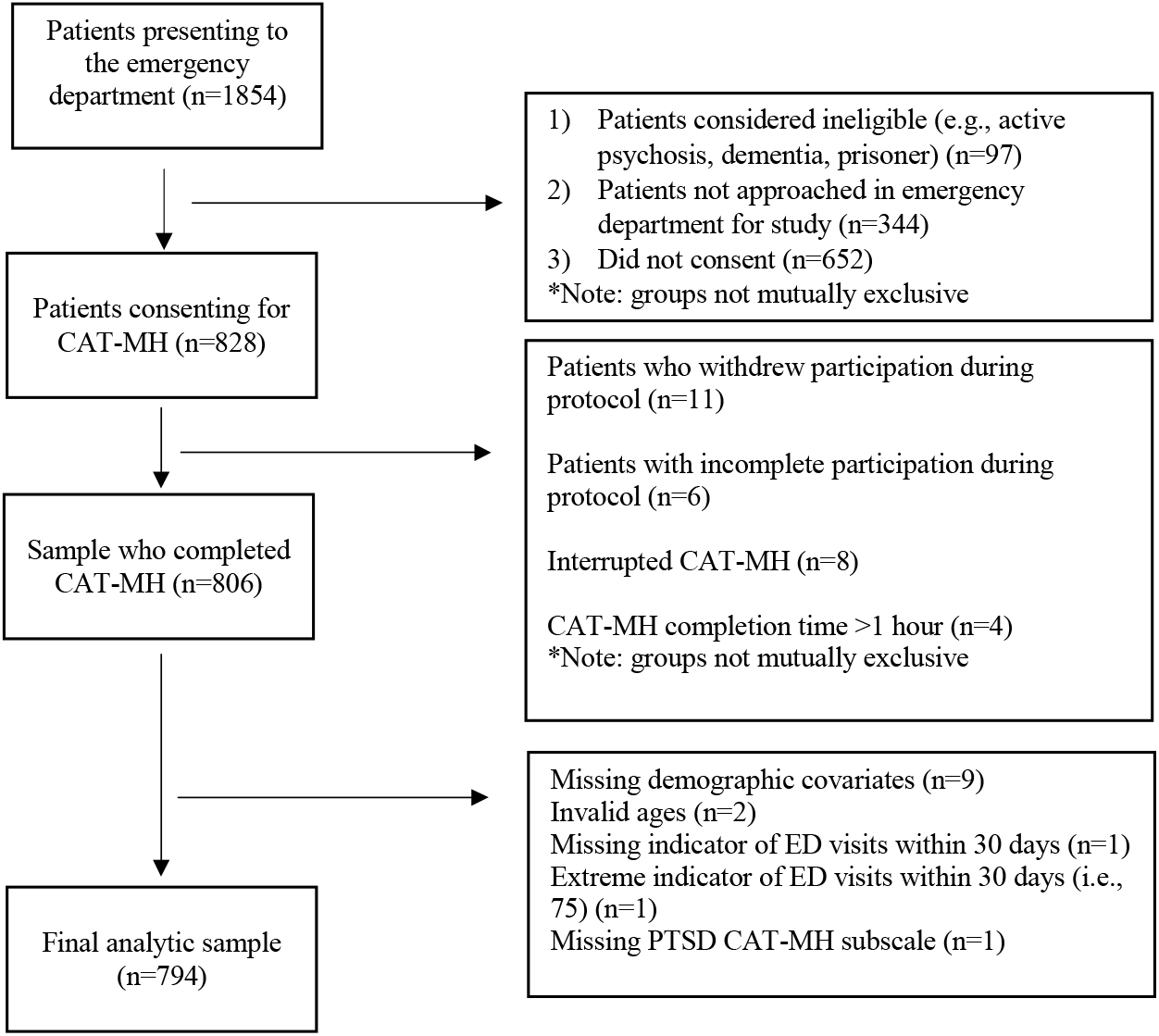
Data flowchart.

**Supplementary Table 1.**
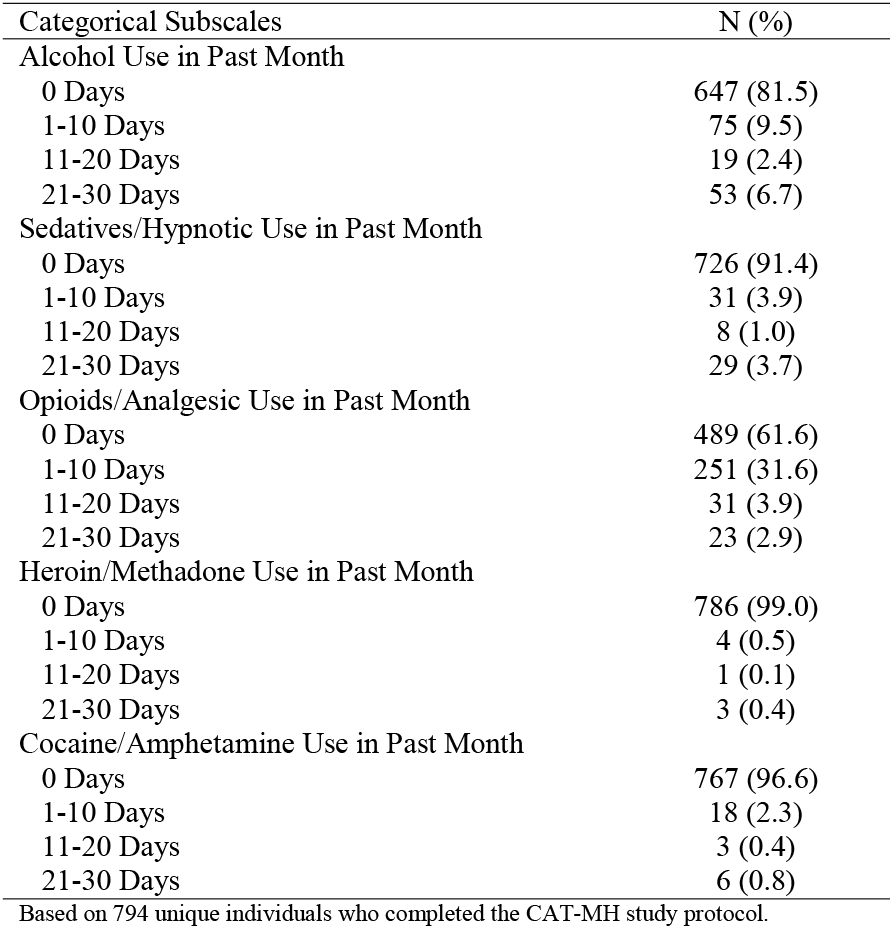
Frequency distribution of CAT-MH specific substance use variables

| Categorical Subscales | N (%) |
| --- | --- |
| Alcohol Use in Past Month |  |
| 0 Days | 647 (81.5) |
| 1-10 Days | 75 (9.5) |
| 11-20 Days | 19 (2.4) |
| 21-30 Days | 53 (6.7) |
| Sedatives/Hypnotic Use in Past Month |  |
| 0 Days | 726 (91.4) |
| 1-10 Days | 31 (3.9) |
| 11-20 Days | 8 (1.0) |
| 21-30 Days | 29 (3.7) |
| Opioids/Analgesic Use in Past Month |  |
| 0 Days | 489 (61.6) |
| 1-10 Days | 251 (31.6) |
| 11-20 Days | 31 (3.9) |
| 21-30 Days | 23 (2.9) |
| Heroin/Methadone Use in Past Month |  |
| 0 Days | 786 (99.0) |
| 1-10 Days | 4 (0.5) |
| 11-20 Days | 1 (0.1) |
| 21-30 Days | 3 (0.4) |
| Cocaine/Amphetamine Use in Past Month |  |
| 0 Days | 767 (96.6) |
| 1-10 Days | 18 (2.3) |
| 11-20 Days | 3 (0.4) |
| 21-30 Days | 6 (0.8) |

Based on 794 unique individuals who completed the CAT-MH study protocol.

**Supplementary Table 2.**
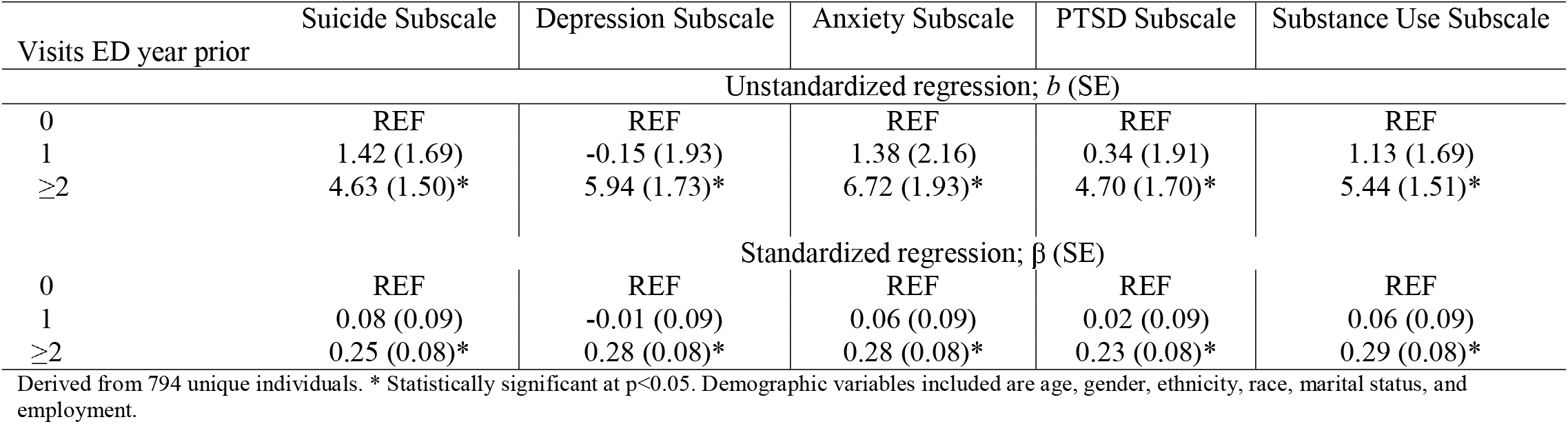
Prediction of CAT-MH from prior year ED visits using linear regression

| Visits ED year prior | Suicide Subscale | Depression Subscale | Anxiety Subscale | PTSD Subscale | Substance Use Subscale |
| --- | --- | --- | --- | --- | --- |
| Unstandardized regression; *b* (SE) |  |  |  |  |  |
| 0 | REF | REF | REF | REF | REF |
| 1 | 1.42 (1.69) | -0.15 (1.93) | 1.38 (2.16) | 0.34 (1.91) | 1.13 (1.69) |
| ≥2 | 4.63 (1.50)* | 5.94 (1.73)* | 6.72 (1.93)* | 4.70 (1.70)* | 5.44 (1.51)* |
| Standardized regression; $\beta$ (SE) | | | | | |
| 0 | REF | REF | REF | REF | REF |
| 1 | 0.08 (0.09) | -0.01 (0.09) | 0.06 (0.09) | 0.02 (0.09) | 0.06 (0.09) |
| ≥2 | 0.25 (0.08)* | 0.28 (0.08)* | 0.28 (0.08)* | 0.23 (0.08)* | 0.29 (0.08)* |

Derived from 794 unique individuals. \* Statistically significant at $p < 0.05$ . Demographic variables included are age, gender, ethnicity, race, marital status, and employment.

**Supplementary Table 3.** Prediction of ED visits within 30 days using logistic regression with continuous predictors

| CAT-MH Subscale | OR (95% CI; p-value)<sup>a</sup> |
| --- | --- |
| Suicide | 1.19 (1.01-1.41; p=0.04) |
| Depression | 1.16 (0.97-1.37; p=0.10) |
| Anxiety | 1.17 (0.99-1.39; p=0.07) |
| PTSD | 1.06 (0.89-1.26; p=0.52) |
| Substance Use | 1.17 (0.97-1.39; p=0.10) |

Derived from 794 unique individuals. ED visits dichotomized into not present/any (0/1). Continuous predictors standardized. Demographic variables included are age, gender, ethnicity, race, marital status, and employment.

**Supplementary Table 4.** Prediction of ED visits in prior year from CAT-MH subscales using ordinal logistic regression

|  | OR (95% CI; p-value)<br>ED visits within 30 days<sup>a</sup> |
| --- | --- |
| Suicide |  |
| Low Risk | REF |
| Intermediate Risk | 1.34 (0.98-1.83; p=0.07) |
| High Risk | 1.59 (0.50-5.05; p=0.43) |
| Depression |  |
| Normal | REF |
| Mild | 1.49 (1.13-1.97; p=0.01) |
| Moderate | 1.33 (0.69-2.53; p=0.39) |
| Severe | 2.04 (1.04-4.02; p=0.04) |
| Anxiety |  |
| Normal | REF |
| Mild | 1.59 (1.11-2.28; p=0.01) |
| Moderate | 1.52 (0.95-2.42; p=0.08) |
| Severe | 1.59 (0.97-2.60; p=0.07) |
| PTSD |  |
| No Evidence | REF |
| Possible | 1.99 (1.27-3.13; p=0.003) |
| Highly Likely | 2.58 (1.20-5.56; p=0.02) |
| Substance Use |  |
| Low Risk | REF |
| Intermediate Risk | 1.35 (0.96-1.90; p=0.08) |
| High Risk | 1.81 (0.77-4.25; p=0.17) |

Derived from 794 unique individuals. Demographic variables included are age, gender, ethnicity, race, marital status, and employment. <sup>a</sup>Ordinal outcome categories included 0, 1, 2, and $\geq 3$ .

